# SOLO study: A single-pill combination strategy in general practice to optimize blood pressure control in a multi-ethnic community

**DOI:** 10.64898/2026.02.24.26346976

**Authors:** R.E. Harskamp, W.R. Naaktgeboren, J. Strijp, Sanne Smits, J.C.L. Himmelreich

## Abstract

**Background:** Hypertension is a major modifiable risk factor for cardiovascular disease, yet blood pressure (BP) control remain suboptimal, particularly in socially disadvantaged communities. Guidelines recommend initiating single-pill combination (SPC) therapy to improve adherence and BP control, but uptake in primary care is limited.

**Objectives:** To evaluate the SOLO care improvement project, promoting SPC initiation among general practitioners (GPs) in Amsterdam Zuidoost, a disadvantaged, multi-ethnic community in The Netherlands with a high hypertension burden.

**Methods:** In a cluster quasi-randomized cluster design, adult hypertensive patients from nine general practices within one health facility were allocated to intervention (IC; n=5) or usual care (UC; n=4). Intervention practices received case-specific guidance on SPC therapy. Outcomes were SPC uptake, changes in systolic and diastolic BP (SBP and DBP), target BP achievement and cardiovascular events. Analyses used intention-to-treat adjusted regression and Cox models, with additional as-treated analysis among SPC users.

**Results:** Among 438 patients (mean age 64.5±12.2 years; median follow-up of 367 days [213-467]), SPC initiation was higher in the IC than US (25.1% vs. 9.6%, p<0.001). SBP/DBP decreased by -15.7/-6.9 mmHg in the IC and -10.4/-4.6 mmHg in the UC. Target BP was more often achieved in the IC (57.3% vs. 48.1%; OR: 1.4, 95%CI:1.0-2.1). Among SPC users, SBP/DBP decreased by -22.4/-10.5 mmHg.

**Conclusion:** Promoting SPC therapy improved blood pressure control, supporting local, targeted implementation as a pragmatic strategy to enhance hypertension management.

**Summary box, bullet points:**

- In the SOLO care improvement project, SPC initiation was increased and improved blood pressure control in routine primary care.
- The real-world implementation and cluster-based comparison enhanced practical relevance and reduced contamination between practices.
- Although conducted in a large community health center, generalizability cannot be assumed; the non-blinded, non-randomized design allows residual confounding.

## Introduction

Hypertension is a major modifiable risk factor for cardiovascular disease (CVD), with approximately 1.4 billion adult patients affected globally, particularly in low- and middle-income countries [1,2]. In the Netherlands, about 2.8 million individuals are treated for hypertension [3]. From these patients, only 21% achieves adequate blood pressure usual care with lower success rates in socially-disadvantaged areas [3,4]. Hypertension contributes significantly to healthcare costs, including 387 million euros in the Netherlands in 2019 [5]. Effective blood pressure management through lifestyle changes and pharmacological treatment is essential for preventing complications and thus improving (long-term) health outcomes. Nevertheless, a substantial proportion of patients continues to have uncontrolled hypertension due to a variety of factors, including poor medication adherence and, especially in primary care, therapeutic inertia [4].

Most guidelines, including the European Society of Cardiology / European Society of Hypertension guideline [6] and the Dutch guideline for primary care physicians [7], currently recommend initiating treatment with more than one hypertensive agent. In this context, single-pill combinations (SPCs), as opposed to multiple pills in free combination, are promoted to reduce complexity and thus improve medication adherence. Indeed, in most studies, SPC therapy has been associated with better medication adherence and blood pressure outcomes compared to free dose regimens [8–12]. Also, some studies indicate that SPC therapy might reduce cardiovascular events and lower overall healthcare expenditure, with no significant increase in adverse events [13–15]. However, despite guidelines recommendations, the real-world use of SPCs in primary care remains modest. Contributing factors include limited physician experience with SPCs or existing concerns that initiating combination therapy is too aggressive and increases side-effects [16].

In response, the SOLO care improvement project was launched in Amsterdam Zuidoost, the Netherlands, a multi-ethnic community with a high hypertensive burden. The aim of the SOLO initiative was to promote SPC initiation among general practitioners (GPs). Here, we evaluate the outcomes of the SOLO care project’s implementation, and discuss the steps required to advance this approach further in primary care.

## Methods

### Study design and setting

The SOLO project was designed as a quasi-experimental, parallel-cluster study, conducted within healthcare facility ‘Gezondheidscentrum Holendrecht’. This facility comprises nine different GP clinics and is located in Amsterdam Zuidoost, the Netherlands, a socially disadvantaged, multi-ethnic community with a high hypertensive burden. The project ran for two years, beginning on November 1, 2023 and ending on October 31, 2025.

The GP clinics within the health facility were assigned to either the intervention or usual care cluster, based on their fixed order of affiliation within the center. The first five GP clinics were allocated to the intervention cluster, and the remaining four served as the usual care cluster. In the intervention cluster, GPs were advised to start SPC therapy in patients with primary hypertension, defined as systolic blood pressure (SBP) ≥140mmg or diastolic blood pressure (DBP) ≥90 mm Hg. The SPC protocol (Appendix A) was made in co-creation with the local pharmacist based on availability, population characteristics, side effects profile, and costs, and consisted of three intensity levels:

- Low: amlodipine/valsartan (5/80-160mg) or valsartan/hydrochlorothiazide (80-160/12.5mg).
- Medium: amlodipine/valsartan/hydrochlorothiazide (5/160/12.5mg).
- High: amlodipine/valsartan/hydrochlorothiazide (10/320/25mg).

In addition to these basic formularies, the protocol advised GPs to individualize antihypertensive treatment based on patient-specific factors, including medical history and ethnicity, preferences and clinical considerations. Moreover, the community pharmacy made sure medications were provided and changed combinations, when not available. All prescribed medications were on-label and reimbursed. The protocol allowed for adjustments to therapy if blood pressure remained insufficiently controlled. GPs and practice nurses were trained on the new study protocol through team meetings and individualized sessions, and the study was conducted in collaboration with the local pharmacist. GPs in the usual care cluster were informed on the option of prescribing SPCs during an information session, but did not receive specific recommendation support and continued to provide care as usual.

### Study population and parameters

Patients were eligible if they were ≥18 years of age, and had a documented diagnosis of hypertension, identified by the ICPC codes K85, K86 and K87 (‘elevated blood pressure’, ‘uncomplicated hypertension’ and ‘hypertension with involvement of target organs’, respectively), between November 1, 2023, and October 31, 2025. Patients were excluded in case of (i) major CVD at baseline, (ii) no follow-up blood pressure measurements or a follow-up duration less than seven days, and (iii) those who received cardiovascular care by a medical specialist in the hospital.

All data were retrieved from the electronic health record systems of the participating GPs. Blood pressures measurements (SBP and DBP) were collected at baseline and follow-up, with the latter being defined as the most recent available measurement occurring at least seven days from baseline. For consistency purposes, analyses were limited to office measurements only, thus not including 30-minutes or 24-hours ambulatory blood pressure recordings. Antihypertensive treatment was documented at baseline, if present, and during follow-up. In cases of discontinuation of SPC, reasons for stopping were recorded. Hypertension-related healthcare visits were defined as both in-person consult and phone calls, and were annualized according to each patients’ follow-up duration. Information on incident CVD, i.e. stroke or myocardial infarction, or death were extracted along with the corresponding date of event.

### Statistical analysis

Patient characteristics were expressed as mean ± standard deviation (SD), median (25^th^ and 75^th^ percentiles [Q1-Q3], or frequencies (percentages), as appropriate. Intention-to-treat linear regression models were used, with cluster allocation (intervention versus usual care) and changes in mean SBP or DBP, as independent and dependent variables, respectively. All models were adjusted for age and sex. An as-treated analysis was conducted among patients who newly started SPC therapy, irrespective of cluster allocation. In a sensitivity analysis, we excluded SPC users at baseline to isolate the potential treatment effect of initiation of SPC therapy. Potential effect modification by older age (>50 years) or sex was explored by adding an interaction term of the fully adjusted model, in which we considered a p value >0.10 as evidence for interaction.

The association between cluster allocation and a combined endpoint of death and incident major CVD, defined as myocardial infarction, cerebrovascular event or heart failure, was evaluated with Cox proportional hazard models, adjusting for the same covariables as the linear regression models. All analyses were performed using R and Rstudio software (version 4.3.3, Rstudio Inc., Boston, MA).

#### Ethics

The Medical Ethics Committee at the Amsterdam UMC reviewed the study protocol (reference W23_07#23.097) and decided that this research does not fall under the scope of the Dutch Medical Research Involving Human Subjects Act (WMO) and granted a waiver for official approval.

## Results

In total, 7,699 individuals were registered in the participating practices, of whom 1,476 had a diagnosis of hypertension. Of these, 941 were considered ineligible due to co-existing recorded CVD (n=465) and adequately controlled blood pressure (n=476). Among the remaining 535 patients, an additional 157 patients were excluded for the following reasons: lack of adequate follow-up (n=65), hypertension management provided by specialist (n=12), and data inconsistencies, including incorrect baseline dates (n=4), uncoded CVD (n=8), and pre-coding errors (n=6). This resulted in a final sample of 440 patients.

Patient characteristics are provided in *Table 1*, stratified by allocation to the intervention cluster (n=199) or the usual care cluster (n=239). Participants were comparable across most baseline characteristics. Average BMI was slightly lower in the intervention compared to the usual care cluster; 28.7 ± 5.6 kg/m^2^ versus 30.4 ± 6.2 kg/m^2^, respectively. Follow-up duration was longer in the intervention cluster (379 days [214-486]) than in the usual care cluster (363 days [211-452]).

**Table 1:** Patient characteristics.

|  | Usual care | Intervention cluster |
| --- | --- | --- |
|  | n=239 | n=199 |
| Age (mean (SD)) | 64.3 (12.12) | 64.7 (12.32) |
| BMI (mean (SD)) | 30.4 (6.18) | 28.7 (5.62) |
| Sex = men (%) | 84 (35.1) | 67 (33.7) |
| Smoking (%) |  |  |
| never | 114 (50.7) | 101 (55.2) |
| former | 56 (24.9) | 47 (25.7) |
| current | 35 (15.6) | 28 (15.3) |
| unknown | 20 ( 8.9) | 7 ( 3.8) |
| Ethnicity (%) |  |  |
| Unknown | 89 (37.2) | 59 (29.6) |
| Dutch | 21 ( 8.8) | 29 (14.6) |
| Ghanaian | 25 (10.5) | 20 (10.1) |
| Surinamese | 85 (35.6) | 68 (34.2) |
| Morrocan | 1 ( 0.4) | 6 ( 3.0) |
| South Asian | 3 ( 1.3) | 6 ( 3.0) |
| Other | 15 ( 6.3) | 11 ( 5.5) |
| Antihypertensive therapy |  |  |
| Beta-blocker | 46 (19.2) | 41 (20.6) |
| Calcium channel blockers | 108 (45.2) | 72 (36.2) |
| ACE inhibitor | 115 (48.1) | 93 (46.7) |
| Diuretics | 73 (30.5) | 83 (41.7) |
| Others | 0 ( 0.0) | 5 ( 2.5) |
| Comorbidities (%) |  |  |
| None | 53 (22.2) | 46 (23.1) |
| CVD | 33 (13.8) | 18 ( 9.0) |
| Diabetes mellitus | 78 (32.6) | 64 (32.2) |
| Hypercholesterolemia | 50 (20.9) | 30 (15.1) |
| Renal impairment | 33 (13.8) | 21 (10.6) |
| Pulmonary disease | 17 ( 7.1) | 25 (12.6) |
| Other | 63 (26.4) | 40 (20.1) |

### SPC uptake

Details on the SPC therapy use are presented in *Table 2*. At baseline, 13.1% (n=26/199) of participants in the intervention cluster and 9.2% (n=22/239) in the usual care cluster used SPC agents. At follow-up, SPC use increased to 25.1% (n=50/199) in the intervention cluster, and remained stable 10.0% (n=24/239) in the usual care cluster (p <0.001 for between-group difference). The median number of consultations was 4 consultations [3-6] in both clusters.

**Table 2:**
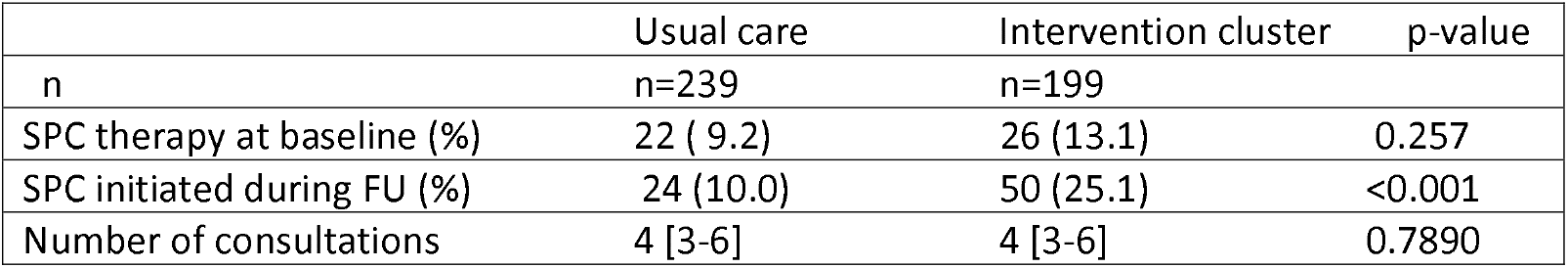
SPC uptake and consultations during follow-up (369 days, 213-467)

|  | Usual care | Intervention cluster | p-value |
| --- | --- | --- | --- |
| n | n=239 | n=199 |  |
| SPC therapy at baseline (%) | 22 (9.2) | 26 (13.1) | 0.257 |
| SPC initiated during FU (%) | 24 (10.0) | 50 (25.1) | <0.001 |
| Number of consultations | 4 [3-6] | 4 [3-6] | 0.7890 |

### Blood pressure outcomes

In both clusters, mean SBP and DBP decreased from baseline to follow-up. In the intervention cluster, the mean reductions in SBP and DBP were -15.7 mm Hg (95% - 18.4; -13.0) and -6.9 mm Hg (-8.4; -5.3), respectively (*Table 3*). In the usual care cluster, the corresponding reductions were -10.4 mm Hg (95% -12.9; -7.8) and -4.6 mm Hg (-6.1; -3.2). Compared to usual care, SBP and DBP decreased significantly more in the intervention cluster (delta -5.3 mmHg (95% -9.0; -1.6) and -2.3 mm Hg (95% -4.4; 0.2), respectively). The proportion of patients who reached their target blood pressure increased in both clusters, with 57.3% in the intervention cluster and 48.1% in the usual care cluster achieving their targets at follow-up. Patients in the intervention were more likely to achieve their target values; OR 1.4 (95%CI 1.0; 2.1).

**Table 3.** Blood pressure outcomes on an intention-to-treat basis.

| **Outcome** | **Cluster** | **Baseline** | **Within-group difference** | **Between-group difference** |
| --- | --- | --- | --- | --- |
|  |  | **mean (SD)** | **delta (95%CI)** | **delta (95% CI)** |
| Systolic | usual care | 155.3 (15.9) | -10.4 (-12.9; -7.8) | reference |
|  | intervention | 156.5 (17.7) | -15.7 (-18.4; -13.0) | -5.3 (-9.0; -1.6) |
| Diastolic | usual care | 90.7 (11.0) | -4.6 (-6.1; -3.2) | reference |
|  | intervention | 91.3 (10.7) | -6.9 (-8.4; -5.3) | -2.3 (-4.4; -0.2) |

### SPC therapy

SPC therapy was initiated in 74 patients in our study (n=74/440; 16.8%), of whom 51 were originally allocated to the intervention cluster. Patients newly started on SPC therapy tended to be younger, non-smoking males with a longer duration of follow-up (415 days [282-531] in SPC group versus 364 days [207-452] in the non-SPC group) (*Supplemental Table S1*).

In the SPC group, SBP decreased by 22.4 mm Hg (95%CI -27.8; -17.1), and DBP decreased by 10.8 mm Hg (95%CI -12.7; -8.9) over time (*Table 4*). In the non-SPC group, the reductions in SBP and DBP were -10.9 mm Hg (95%CI -12.7; -8.9) and - 10.5 mm Hg (95%CI -13.5; -7.5), respectively. Compared to non-SPC users, initiation of SPC therapy corresponded with a greater reduction in both SBP (delta - 12.2 mm Hg (95%CI -17.0; -7.2) and DBP (delta -5.5 mm Hg (95%CI -8.3; -2.7)) compared with patients who did not start SPC treatment. Patients with new SPC treatment achieved more often their target blood pressure values: OR 1.9 (95% CI 1.1; 3.2). The sensitivity analysis, in which we excluded patients who were using SPC at baseline, did not change the overall conclusion (*Supplemental Table S2*). Also, we found no evidence for interaction by older age or sex (p>0.10 for both).

**Table 4.** Blood pressure outcomes on an as-treated analysis among patients who started on SPC therapy.

| Outcome | Cluster | Baseline | Within-group difference | Between-group difference |
| --- | --- | --- | --- | --- |
|  |  | mean (SD) | delta (95%CI) | delta (95% CI) |
| Systolic | No SPC | 154.4 (15.3) | -10.8 (-12.7; -8.9) | reference |
|  | SPC users | 163.1 (21.0) | -22.4 (-27.8; -17.1) | -12.1 (-17.0; - 7.2) |
| Diastolic | No SPC | 90.6 (10.5) | -4.7 (-5.8; -3.5) | reference |
|  | SPC users | 92.9 (12.3) | -10.5 (-13.5; -7.5) | -5.5 (-8.3; -2.7) |

### Cardiovascular events during follow-up

In total, six patients experienced a major cardiovascular event during registered follow-up (cerebrovascular events: n=4 and myocardial infarction: n=2). No deaths were recorded. The occurrence of a major cardiovascular event was not associated with allocation to either cluster nor with the initiation of SPC treatment (hazard ratio 1.08 (95% CI 0.22; 5.35) and hazard ratio 0.69 (95% CI 0.08; 6.02), respectively).

## Discussion

The SOLO care improvement project was associated with a higher uptake of SPC therapy and improved blood pressure control in a socially disadvantaged community in the Netherlands with a high hypertension burden. These results support that local SPC implementation projects could lead to more effective hypertension management in primary care.

### Previous studies

Our findings of increased SPC initiation and better blood pressure outcomes are in line with a substantial body of evidence supporting the effectiveness of SPC therapy in hypertension management. The efficacy of SPC therapy in terms of medication adherence with subsequent blood pressure control has been emphasized in the current hypertension guidelines, including the Dutch primary care hypertension guideline and the 2024 ESC hypertension guideline [6,17]. Four recent meta-analyses have consistently demonstrated that initiating SPC therapy is associated with greater reductions in SBP and DBP compared with monotherapy, free-equivalent combinations, or usual care [11,18–20]. Additionally, SPC therapy is associated with a higher proportion of patients reaching their blood pressure value, with control rates up to 80% among SPC patients versus 46-65% for alternative regimens[8,18,21]. Notably, these findings were observed across diverse population, including those with therapy-resistant hypertension or low-to-middle-income countries.

Our study differs from most existing literature in its real-world implementation setting. Whereas most studies compared SPC therapy to non-SPC regimens within the context of a clinical trial, our study evaluated the broader impact of a routine care-based improvement initiative designed to promote SPC initiation among GPs. In this context, some patients already used a SPC at baseline (11%) and ‘only’ a quarter of the patients in the intervention cluster initiated SPC therapy. Although this likely dilutes the anticipated treatment effect, we were still able to observe a substantial reduction in SBP and DBP which, which in direction and magnitude, aligns with clinical studies[8,18,21]. In other words, the findings from SOLO support the notion that the benefits of SPC therapy can translate to less-controlled, real-world conditions, or at least to a Dutch primary care context. Overarching, given the low SPC uptake observed in the usual care cluster, these findings underscore the potential value of other local, targeted care implementation initiatives.

### Clinical relevance of BP reductions

The observed reduction in SBP and DBP (-5.3 mmHg and -2.3 mmHg in the intention-to-treat analysis, respectively) are modest at an individual level. Yet, on a population level, even small decreases in blood pressure can translate to a meaningful reduction in cardiovascular disease risk. A large meta-analysis reported that a 5 mm Hg reduction in SBP reduced the risk of incident major cardiovascular events by nearly 10%, regardless of previous cardiovascular disease, and even among individuals with normal or high-normal blood pressure values [22].

Nevertheless, the magnitude of the blood pressure reduction that can be expected depends on pretreatment SBP, often referred to as the Wilders principle: pre-treatment value determines post-treatment response. This principle may partly explain the large decrease in SBP and DBP observed in the as-treated analysis among SPC users in our study, whose baseline values were 163 mm Hg and 93 mm Hg, respectively. Nonetheless, when limiting our analyses to those that newly started on SPC therapy (thereby excluding the baseline SPC users), we observed comparable point estimates, yet with wider confidence intervals, supportive of a therapeutic effect of SPC initiation. This aligns with findings of clinical studies, in which SPC agents were found to be more potent among individuals with higher pretreatment values, with smaller absolute reductions in those with mildly elevated blood pressure. Of importance, SPC therapy was not associated with an increased rate of adverse events, including those related to hypotension across multiple meta-analyses [11,18,19]. This finding is relevant in the context of a qualitative study among Dutch GPs, which reported that interviewed GPs preferred a stepped monotherapy approach to better identify potential side effects [23]. The low uptake of SPC in the usual care cluster in our study, with only 11% of the patients using SPC, suggests substantial room for improvement in route primary care practices. This is particularly true for the context in which the study was conducted, a socially disadvantaged community with a high hypertension burden. Low socioeconomic status is associated with poor medication adherence, due to, among others, low health literacy and complex treatment regimens [24,25]. This contributes to persistently uncontrolled hypertension and larges health inequities. To these individuals, SPC therapy may be particularly valuable, given that SPC therapy is known to simplify treatment and improve adherence by reducing pill burden. In this context, SPC therapy may be especially effective in improving medication adherence in such populations and thus may help addressing health inequities in hypertension management.

### Strengths and limitations

This study was rigorously conducted, in a real-world setting, with clear guidance/protocol and consistent use of office blood pressure readings. However, this study also has several limitations that should be considered when interpreting the findings. The non-randomized allocation of the intervention allows for residual confounding, including differences between general practice clinics and potential selection within patient flow. In addition, the follow-up period was relatively short, limiting the ability to detect cardiovascular events. Finally, as the study was conducted within a single healthcare facility in Amsterdam Zuidoost, the generalizability of the results to other primary care settings may be limited. At the same time, the pragmatic design and embedding within routine primary care represent important strengths, enhancing the relevance of the findings for real-world clinical practice.

### Future directions

Future research should move beyond demonstrating short-term blood pressure reductions toward understanding how effective strategies can be sustainably implemented at scale in community settings. While there is strong evidence that early and intensive blood pressure lowering improves control rates, important gaps remain regarding long-term clinical outcomes, cost-effectiveness, and equity across diverse populations. In addition, research should focus on identifying which patients benefit most from simplified treatment strategies and how treatment intensity can be tailored to baseline risk without increasing adverse effects. Finally, implementation-focused and qualitative studies are essential to address barriers at the patient, clinician, and health system level. This is particularly relevant in multi-ethnic and socioeconomically diverse communities, where hypertension burden is highest and control rates lowest.

## Conclusion

In a socially disadvantaged, multi-ethnic primary care setting, promoting SPC therapy increased its initiation and led to greater blood pressure reductions and improved target blood pressure achievement compared with usual care. These findings support SPC promotion as a pragmatic strategy to improve hypertension management in community practice.

## Data Availability

All data produced in the present work are contained in the manuscript.

