## Supplemental Tables 1 & 2 for "SOLO study: A single-pill combination strategy in general practice to optimize blood pressure control in a multi-ethnic community"

**Supplemental Table S1.** Characteristics of patients that started on SPC therapy compared to those who did not started with SPC

no SPC	started SPC

n	364	74

intervention cluster (%)	149 (40.9)	50 (67.6)

age (mean (SD))	65.00 (12.16)	62.03 (12.16)

bmi (mean (SD))	29.91 (5.99)	28.21 (5.87)

sex = men (%)	119 (32.7)	32 (43.2)

smoking (%)

never	172 (50.7)	43 (62.3)

former	88 (26.0)	15 (21.7)

current	57 (16.8)	6 ( 8.7)

unknown	22 ( 6.5)	5 ( 7.2)

ethnicity (%)

unknown	128 (35.2)	20 (27.0)

dutch	38 (10.4)	12 (16.2)

ghanaian	34 ( 9.3)	11 (14.9)

surinamese	128 (35.2)	25 (33.8)

morrocan	6 ( 1.6)	1 ( 1.4)

indian	9 ( 2.5)	0 ( 0.0)

other	21 ( 5.8)	5 ( 6.8)

hypertension (%)	314 (86.3)	61 (82.4)

Antihypertensive medication

Beta-blocker (%)	72 (19.8)	15 (20.3)

Calcium channel blocker	153 (42.0)	27 (36.5)

Renin-angiotension blocker	175 (48.1)	33 (44.6)

Diuretics	125 (34.3)	31 (41.9)

comorbordities (%)

none	74 (20.3)	25 (33.7)

CVD (%)	46 (12.6)	5 ( 6.8)

diabetes mellitus(%)	122 (33.5)	20 (27.0)

hypercholesterolemia (%)	75 (20.6)	5 ( 6.8)

renal impairment (%)	46 (12.6)	8 (10.8)

pulmonary disease (%)	32 ( 8.8)	10 (13.5)

Follow-up time (days)	364 [207-452]	415 [282-531]

**Supplemental Table S2.** Sensitivity analysis without SPC users at baseline (n=48)

**Outcome	Cluster	n=	Baseline	Within-group	Between-group**

**difference	difference**

**mean (SD)	delta (95%CI)	delta (95% CI)**

Intention-to-treat

Systolic	usual care	217	155.4 (15.9)	-10.8 (-13.5; -8.1)	reference

intervention	173	156.6 (17.6)	-16.0 (-19.0; -13.0)	-5.2 (-9.2; -1.2)

Diastolic	usual care	217	913 (11.0)	-5.1 (-6.6; -3.5)	reference

intervention	173	91.1 (10.5)	-6.6 (-8.3; -4.9)	-1.7 (-4.0; 0.5)

**n baseline (%)	n follow-up (%)	odds ratio (95%CI)**

On target*	usual care	217	44 (20.3)	106 (48.8)	reference

Intervention	173	34 (19.7)	100 (57.8)	1.4 (0.9; 2.1)

As-treated

Systolic	No SPC	331	154.5 (15.3)	-11.1 (-13.1; -9.1)	reference

SPC users	59	163.6 (21.5)	-24.3 (-30.6; -18.0)	-13.5 (-18.9; -8.0)

Diastolic	No SPC	331	90.9 (10.6)	-4.8 (-6.0; -3.7)	reference

SPC users	59	92.8 (12.1)	-10.9 (-14.4; -7.4)	-5.7 (-8.8; -2.6)

**n baseline (%)	n follow-up (%)	odds ratio (95%CI)**

On target*	No SPC	331	67 (20.2)	165 (49.8)	reference

SPC users	59	11 (18.6)	41 (69.5)	2.5 (1.4; 4.6)

*reaching guideline recommended systolic and diastolic blood pressure targets

.
